# Molecular portrait of prostate tissue probed by MALDI Imaging Mass Spectrometry

**DOI:** 10.1101/2021.03.21.21254051

**Authors:** Surya Kant Choubey, Amrita Mitra, Rajdeep Das, Pritilata Rout, Amit Kumar Mandal

## Abstract

Benign prostatic hyperplasia (BPH) is the most common condition in aging men, associated with lower urinary tract symptoms. BPH has been suggested to be a risk factor for certain urologic cancers, but the current evidence is inconsistent. The gold-standard method for the diagnosis of BPH is histopathology. Histopathology displays the cellular morphologies of tissues wherein characteristic changes pertaining specifically to BPH can be identified. However, the onset of BPH might be associated with minimal phenotypic changes in cellular morphologies in tissues that histopathology might not be able to detect successfully. Therefore, to understand the onset of a disease and its pathogenesis it is important to investigate the detailed molecular profiles associated with the disease that might help in the diagnosis and to understand the insights of the disease pathogenesis. Over the last decade, imaging mass spectrometry has been used to explore the spatial distribution and expression profiles of several molecules with their two-dimensional heterogeneity retained across the tissues. In the present study, using MALDI mass spectrometry based tissue imaging platform, we observed the expression of several proteins across human prostate tissue sections diagnosed with BPH. The proteins were identified and characterized using tissue proteomics approach. We could successfully identify the on-tissue distribution of ribonuclease T2, vinculin, isoform 2 of tropomyosin alpha-3 chain and mitochondrial citrate synthase proteins using this approach. Therefore, imaging mass spectrometry might be a potential tool to complement the findings of histopathology in diagnosis of BPH in human prostate tissues.

## Introduction

Benign prostatic hyperplasia (BPH) is a non-malignant enlargement of the prostate caused by cellular hyperplasia leading to benign tumors. It is a common age-associated disease affecting ∼70% of men aged 70 years or above. BPH can be a bothersome and potentially severe condition. Not only can it lead to lower urinary tract symptoms and diminish patients’ quality of life but it may also be associated with certain male urologic cancers such as prostate cancer and bladder cancer. Initial screening of the enlarged prostate for diagnosis of BPH involves digital rectal examination (DRE) and transurethral ultrasonography (TRUS). DRE is used to screen for palpable tumors in the prostate gland [1]. As it requires manual intervention, it is not always efficient to detect tumors in the prostate gland [2]. In TRUS, an ultrasound probe is used to image the entire prostate gland. This technique is limited to diagnose distantly located tumors in the prostate gland[3]. In the routine diagnosis for presence of tumors in prostate gland, prostate specific antigen (PSA), a peripheral blood biomarker, is quantitatively assayed where the a value of ≤ 4 ng/mL of PSA in blood is considered to be associated with BPH of prostate gland [4] with reported exceptions [5]. Another approach for the diagnosis of BPH is based on the histopathological examination of tissue biopsies.

Imaging mass spectrometry (IMS) has enabled to visualize the spatial distribution of several proteins in a single scan without the perturbation of the two-dimensional heterogeneity of their distribution across a tissue section [6, 7]. In the present study, using MALDI mass spectrometry, we imaged the distribution of proteins of across human prostate tissue sections which were characterized as BPH based on the histopathological examination. The proteins were identified and characterized using proteomics platform.

## Materials and methods

### Materials

Superfrost^™^ microscopic slides were purchased from Thermo Fisher (USA). Analytical grade ethanol, glacial acetic acid, chloroform and red phosphorous were obtained from Merck (Germany). Sequencing grade bovine trypsin, dithiothreitol, iodoacetamide, β-octylglucopyranoside and α-Cyano-4-hydroxycinnamic acid matrix were purchased from Sigma Aldrich (St. Louis, MO). Water and acetonitrile were of LC−MS grade and obtained from Honeywell (California, USA). All other chemicals used were of analytical grade.

### Ethics statement

The study was approved by the Institutional Ethical Committee (IEC approval no. 27/2014), St. John’s Medical College, Bangalore, Karnataka, India. All the procedures were carried out in accordance with the available guidelines and regulations for human participants in research.

#### Selection of subjects

Patients presenting with benign prostatic hyperplasia (BPH) to the Department of Urology at St. John’s Medical College Hospital, Bangalore, were invited to participate in the study. Written consents were obtained from patients prior to collection of samples.

#### Collection and sectioning of prostate tissue samples

Five prostate tissue bits were collected in normal saline suspected with BPH, from a patients undergoing the procedure of transurethral resection of the prostate (TURP). All these tissue bits and core were pathologically diagnosed with BPH. The tissue samples were mounted on microtome chucks and fixed in ice for sectioning in a cryomicrotome. Sections of 8 µm thickness were taken and thaw-mounted on microscopic glass slides. Two serial sections for each tissue were collected, one each for histopathological examination and MALDI imaging experiment. The remaining cores of tissues were desiccated and stored in −80 °C until used for tissue proteomics experiments.

#### Processing of tissue section for MALDI imaging experiment

The slides containing the tissue sections were desiccated for 45 mins. For fixing the tissue sections and removal of lipids and salts, the tissue sections were treated stepwise with different solutions as follows: 70% ethanol (30 seconds), 100% ethanol (30 seconds), 6:3:1 of ethanol:chloroform:glacial acetic acid – Carnoy’s solution (2 minutes), 100% ethanol (30 seconds), 0.2% trifluoroacetic acid in water (30 seconds) and 100% ethanol (30 seconds). Subsequently, the slides with tissue sections were desiccated for 30 minutes to remove all the solvents, followed by *in situ* trypsin digestion and spraying of matrix.

#### Reduction, alkylation, *in situ* trypsin digestion and matrix application

5mM dithiothreitol and 10 mM iodoacetamide solutions were used for reduction and alkylation procedures respectively. Trypsin solution (0.1 mg/ml) in 50 mM NH_4_HCO_3_, pH 7.4 containing 1% β-octylglucopyranoside was used for digestion. All the above solutions were sprayed over the tissue section at the flow rate of 6 μl/min for 6 layers (Suncollect sprayer/spotter, Germany). The slides were desiccated and incubated for 30 minutes on a dry bath maintained at 60 °C after dithiothreitol treatment. Similarly, following iodoacetamide treatment, the slides were incubated for 30 mins in dark. Following trypsin cycles, the slides were desicated and incubated at 37 °C overnight. α-Cyano-4-hydroxycinnamic acid (HCCA) (5 mg/ml in 1:1 ACN/0.5% aqueous TFA) matrix was sprayed for 25 cycles over the tissue sections at the flow rate of 10 μl/min for the first cycle, followed by 20 μl/min for the second cycles, 30 μl/min for the third cycle and 40 μl/min constantly for the rest of the cycles.

#### MALDI MS imaging of tissue sections

The samples were analyzed using a MALDI Q-TOF mass spectrometer calibrated with red phosphorous (Synapt G2S*i*, WATERS, UK). Mass spectra were acquired in positive polarity in the m/z range 1000 to 4500 with a scan time of 1 sec using the analyzer in sensitivity mode. In general, the quadrupole mass analyzer that was used here in the MS scan, does not transmit all ions with the equal efficiency. Rather, it acts as a broad-band filter that allows passage of ions with m/z values from 0.8 × mass 1 (lower mass end). On the high mass end, the transmission of ions occurs for m/z up to 5 × mass 3. To allow equal transmission of ions in the m/z range 1000 to 4500, the following quadrupole ion transmission profile used: mass 1 (m/z 100), dwell time (4%), ramp time from mass 1 to mass 2 (1%), mass 2 (m/z 900), dwell time (90%), ramp time from mass 2 to mass 3 (5%), and mass 3 (m/z 900). The spatial resolution, measured in terms of pixel size, was optimized at 90 µm in both X and Y directions. The laser was operated at a repetition rate of 1000 Hz and with laser energy of 370 kV. The instrument was operated using a trap collision energy of 10 kV and trap cooling nitrogen gas flow rate of 2.5 mL/min. Image acquisition was carried out using High Definition Imaging (HDI) software, v 1.4 coupled to MassLynx v 4.1 (WATERS, UK).

#### Analysis of images using HDI v 1.4 software

The acquired data were analyzed using HDI software (WATERS, UK). One thousand most intense peaks inclusive of their isotopic distributions were selected from the acquired data for processing in HDI software and to match their specific localization on the tissue sections in terms of X and Y coordinates. The parameters used for processing the data were as follows: m/z window = 0.1 Da; MS resolution = 12000; internal lock mass peak = 2272.1; lock mass tolerance = 0.3 amu; lock mass minimum signal intensity = 500 counts. The intensities of all the peaks were normalized by the total ion count (TIC), where the absolute intensities of each peak was divided by the sum of the absolute intensities of all peaks. The normalized values were plotted to obtain the images corresponding to the distribution of multiple peptides on the tissue sections.

### Tissue proteomics of human prostate samples

#### Processing of the collected tissue samples

One core of prostate tissue samples was lyophilized for 4 hours. The lyophilized tissue was crushed to powder. 0.2 mg of the crushed sample was suspended in tissue lysis buffer containing 10 mM Tris HCl, 0.15 M NaCl, 1 mM EDTA in phosphate buffered saline, pH 7.4 with 1% β-octylglucopyranoside. 10 µL of protease inhibitor cocktail was added to the suspension and subsequently sonicated for 30 min with intermittent cooling. After sonication, the solution was incubated for 1 hour at 4 °C with mild agitation. The suspension was centrifuged at 14,700 x g for 45 min and the supernatant was dialyzed overnight with frequent changes against 50 mM NH_4_HCO_3_ buffer, pH 7.4. After dialysis, the protein concentration was measured by Bradford assay method according to the manufacturer’s protocol and followed by in-solution digestion.

#### In-solution digestion of the proteins from tissue samples

Samples were denatured by using 0.2% Rapigest at 80 °C for 15 min. Subsequently, the disulphide linkages of proteins were reduced by 5 mM dithiothreitol for 30 min at 60 °C, followed by alkylation of the reduced –SH groups with 10 mM iodoacetamide for 30 min in dark at room temperature. Modified sequencing grade trypsin, reconstituted in 50 mM NH_4_HCO_3_, pH 7.4, was added to the protein extracts at an enzyme:substrate ratio of 1:10 and incubated overnight at 37 °C. Following this, the samples were acidified in 0.5% (v/v) formic acid (FA) and incubated at 37 °C for 90 mins to facilitate hydrolysis of Rapigest detergent. Samples were then be centrifuged for 45 min at 4000 x g and the supernatant was collected and spiked with 50 fmoles/μl of internal standard yeast enolase (WATERS, UK). Subsequently, the samples were carried forward for nanoLC-ESI-MS analysis.

#### nanoLC fractionation

For tissue proteomics, the samples were analyzed at Proteomics Facility, Centre for Cellular and Molecular Platforms (CCAMP), National Centre for Biological Sciences (NCBS), Bangalore. The proteolytic peptides were fractionated in EASY-nanoLC 1200 UHPLC system (Thermo Scientific, Germany), calibrated using Pierce^™^ LTQ ESI Positive Ion Calibration Solution (Thermo Scientific, Germany). Briefly, 600 ng of total tryptic digest of the experimental sample was loaded directly into a C18 Acclaim^™^PepMap^™^ Rapid Separation Liquid Chromatography column (3 μm, 100 Å ⨯75 μm ⨯ 50 cm). The aqueous phase consisted of water/0.1% FA (solvent A) and the organic phase comprised of acetonitrile/0.1% FA (solvent B). The flow rate of solvents through the column was 300 nL/min. A 90 min run program was designed, starting with a linear gradient of solvent B from 5-40% for 70 min, followed by 40-80% of solvent B for 8 min, and finally 80-95% of solvent B for 12 min. The column temperature was maintained at 40 °C during the run in all experiments. The eluted peptides were analyzed in a mass spectrometer with an orbitrap mass analyser (OrbitrapFusion^™^ mass spectrometer, Thermo Scientific, San Jose, California, USA).

#### Data acquisition

The data were acquired in the mass range of 375−1700 m/z at 120000 FWHM resolution (at 200 m/z). The instrument was operated in positive ion mode with a source temperature of 350 °C, capillary voltage of 2.0 kV. Poly-siloxane (445.12003 m/z), used as lock mass, was continuously infused throughout the acquisition. The instrument was set to run in top speed mode with 3 s cycles for the survey and the MS/MS scans. After a survey scan, tandem MS was performed on the most abundant precursors exhibiting charge states from 2 to 7 with the intensity greater than 1.0e^4^, by isolating them in the quadrupole. Higher-energy collisional dissociation (HCD) was used for fragmentation of the ions where the normalized collision energy was maintained at 35% and resulting fragments were detected using the rapid scan rate in the ion trap. The automatic gain control (AGC) target for MS/MS was set to 4.0e^5^ and the maximum injection time limited to 50 msec. The experimental samples were run in duplicate.

#### Proteomics Data Analysis

The acquired data was processed using Thermo Scientific(tm) Proteome Discoverer(tm) software version 2.1. MS/MS spectra were searched with the SEQUEST^®^ HT engine against human proteome database incorporated with yeast enolase sequence details. During analysis, the parameters used were as follows: proteolytic enzyme, trypsin; missed cleavages allowed, two; minimum peptide length, 6 amino acids. Carbamidomethylation (+57.021 Da) at cysteine residues was kept as a fixed modification. Methionine oxidation (+15.9949 Da) was set as variable modifications. The mass tolerances for precursor and product ions were kept at 10 ppm and 0.6 Da respectively. Peptides were confidently matched to protein sequences with a maximum false discovery rate of 1% as determined by the Percolator^®^ algorithm.Protein groups were further identified with a criteria of having the presence of at least two unique peptides.

## Results and discussion

### Imaging of prostate tissues

The gold standard method for the diagnosis of BPH is histopathology. In this procedure, the aberrances in tissues associated with disease conditions is based on the observation of cellular morphology of hematoxylin and eosin (H & E) stained tissue sections [8]. Using mass spectrometry based imaging platform, we imaged ribonuclease T2, vinculin, isoform 2 of tropomyosin alpha-3 chain and mitochondrial citrate synthase proteins across prostate tissue sections which were diagnosed as BPH based on histopathological examination. Mass spectrometry based tissue imaging involves proteolytic digestion to identify and characterize proteins. However, the number of observed proteolytic peptides and their signal intensities are generally found to be poor. The disulfide linkages in the functionally active structure of proteins in the tissues limits the desorption of proteolytic peptides from the tissue surface which results in reduced number of proteins observed in the recorded image. We developed a method to enhance the observed number of proteolytic peptides by linearizing the complex structures of proteins using *in situ* reduction and alkylation of the disulfide linkages followed by proteolytic digestion. The protocol described above involves use of dithiothreitol and iodoactamide prior to proteolytic digestion with trypsin. Dithiothreitol is a reducing agent which helps to break the disulfide linkages of proteins and thus linearizes the complex structures of the proteins. Iodoacetamide is an alkylating reagent and prevents the re-folding of the linearized protein molecules as the sulfhydryl groups gets chemically modified with the acetamide group in this reaction. Thus, this protocol resulted in increased coverage of the peaks and enhanced signal to noise ratio. The mass spectra obtained for a BPH sample is shown in Figure 1. We observed the distribution of various proteins on the BPH tissue sections, as shown in Table 1.

**Table 1:**
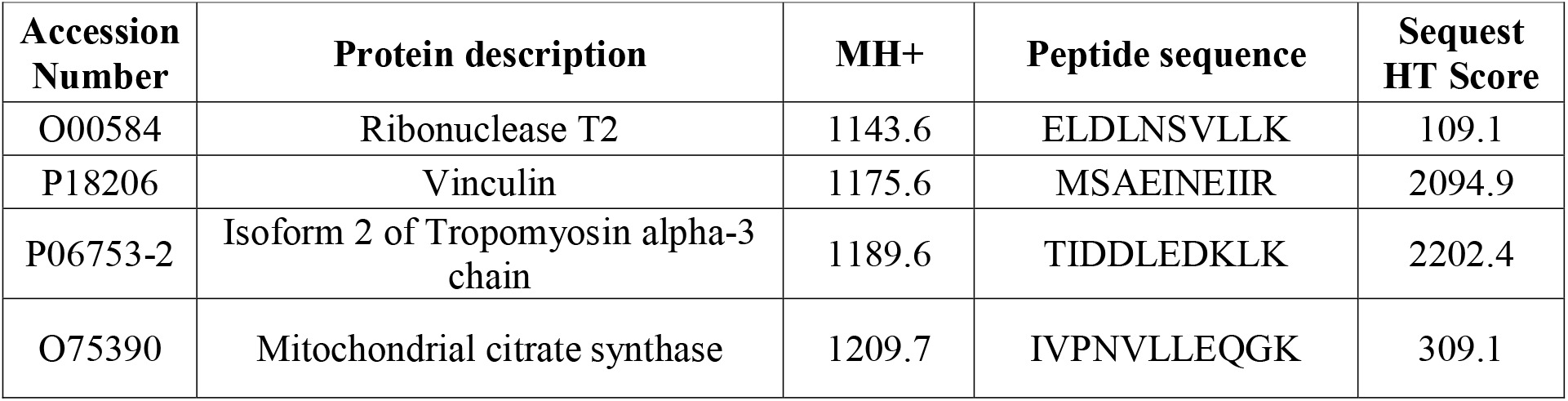
The proteomic characterization of peptides observed in imaging mass spectrometric analysis of BPH prostate tissues Mass spectrometric tissue. images of three representative BPH tissue sectionsare shown in Figures2 and 3. Figures2 and 3 shows the spatial distribution of various analyte ions on a BPH prostate tissue section obtained from TURP procedure.

**Figure 1:**
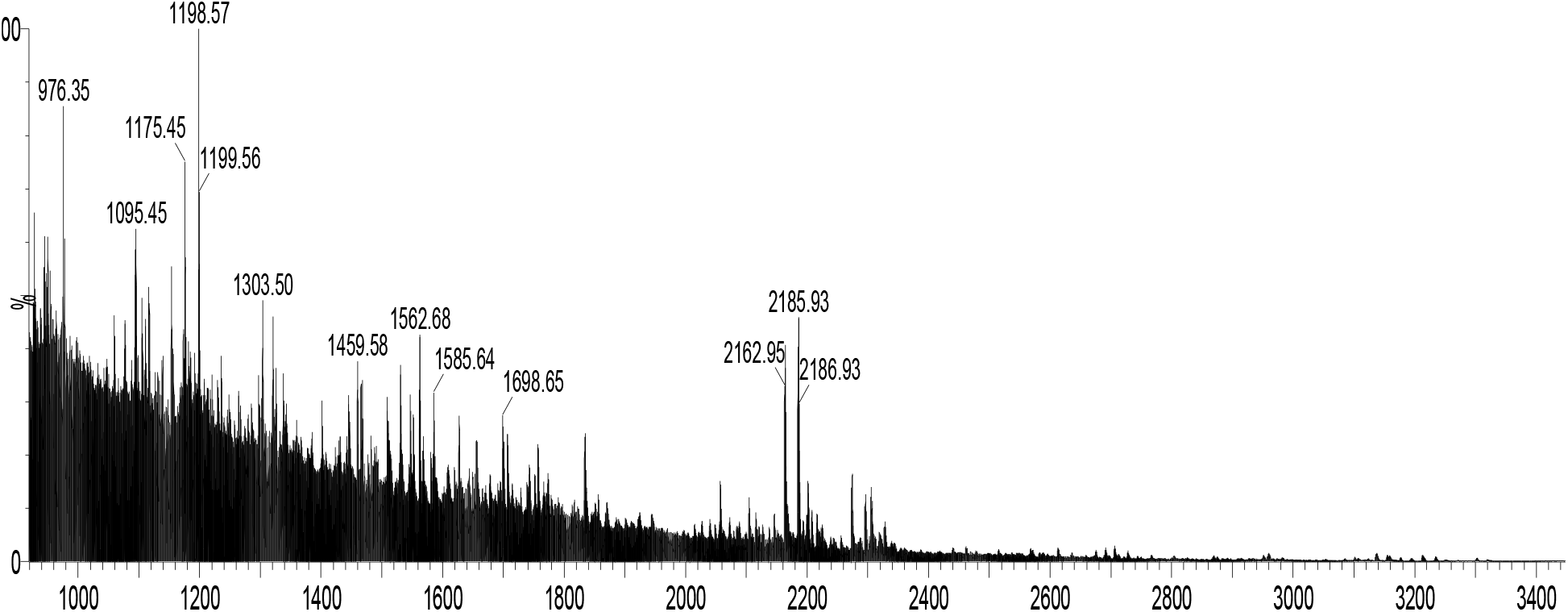
MALDI mass spectra obtained upon imaging of a tissue sample obtained from a patient with benign prostate hyperplasia.

**Figure 2.**
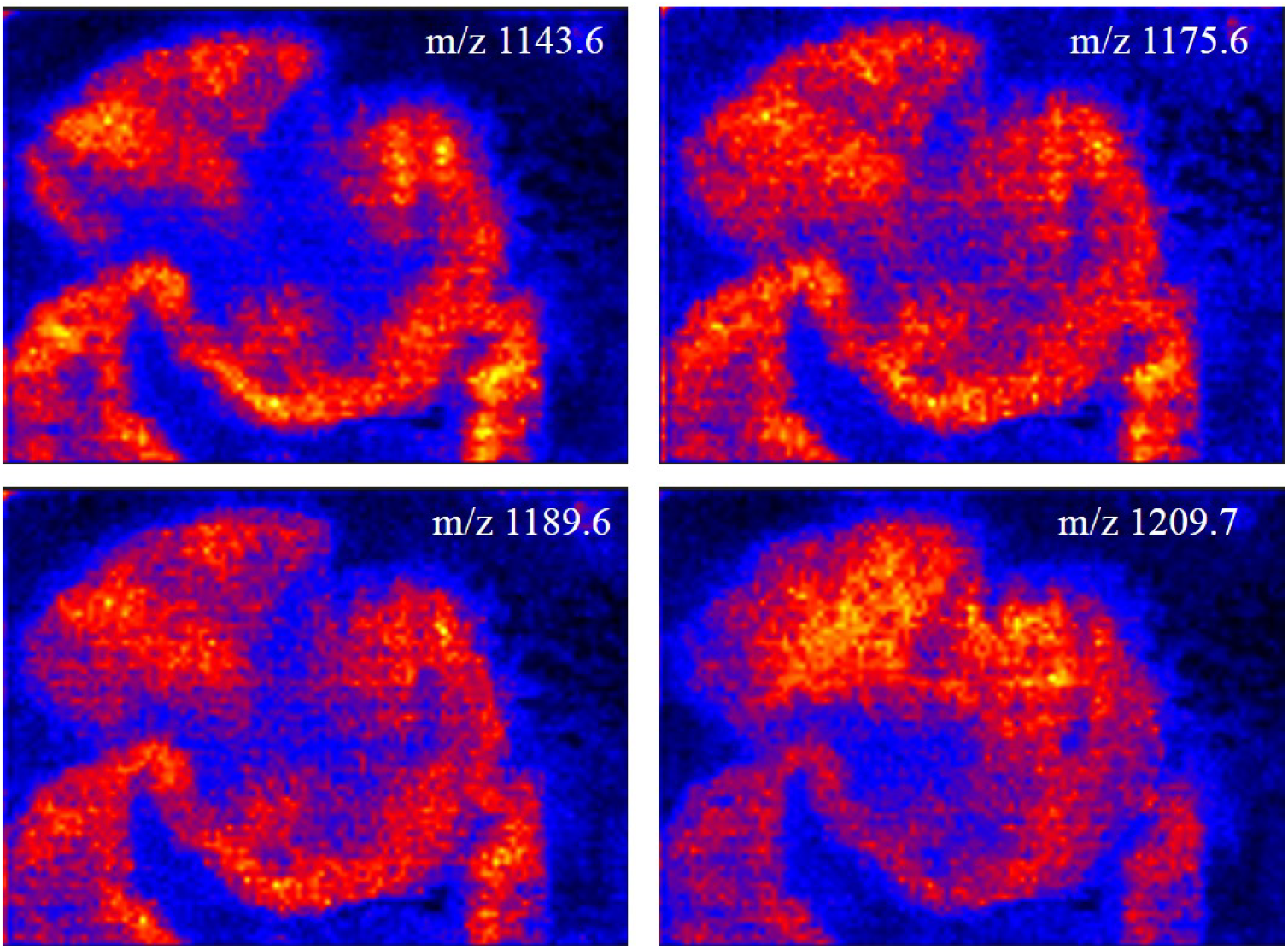
Spatial distribution of peptides with m/z 1143.6, 1175.6, 1189.6 and 1209.7 on a TURP tissue specimen collected from a patient with benign prostate hyperplasia.

**Figure 3:**
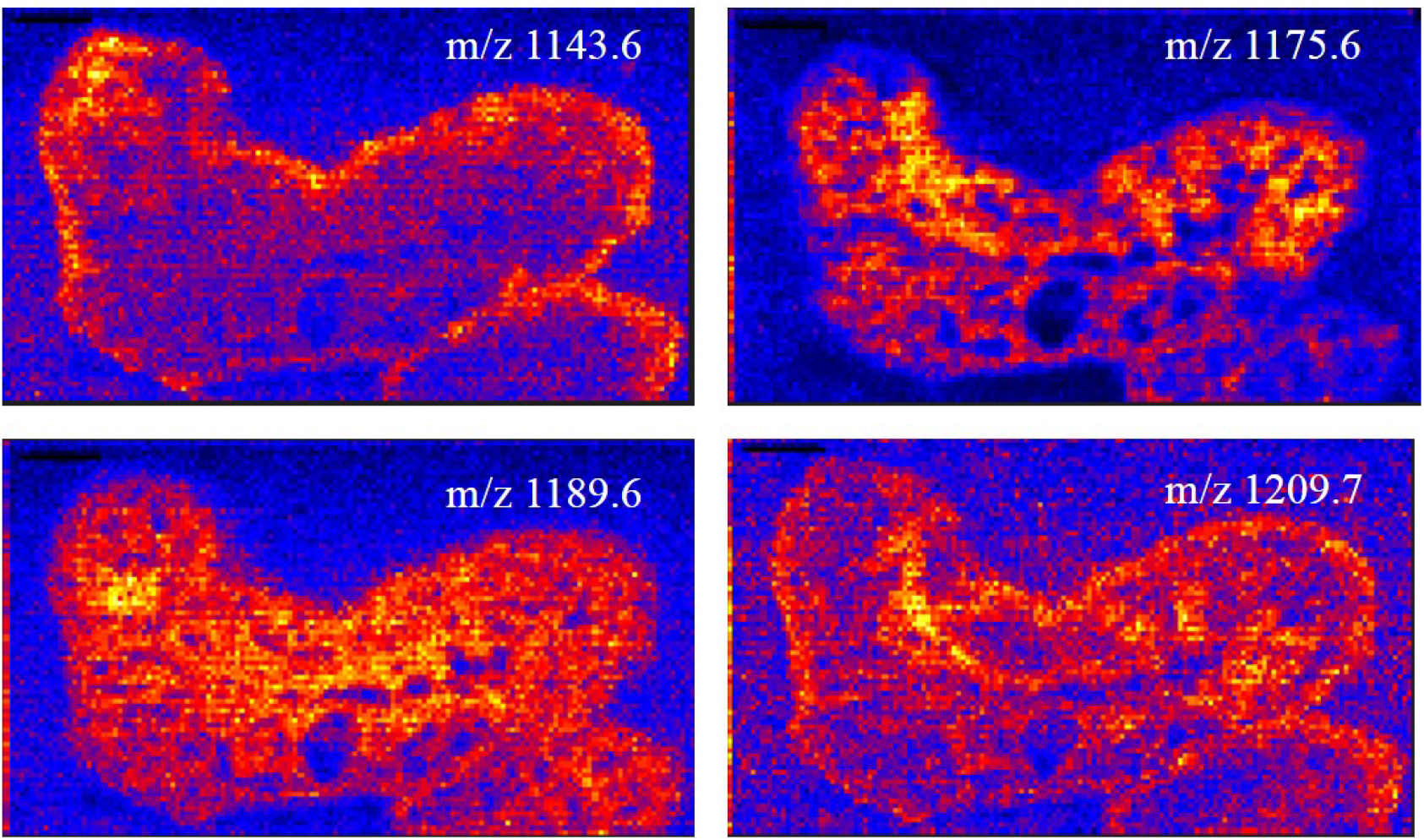
Spatial distribution of peptides with m/z 1143.6, 1175.6, 1189.6 and 1209.7on a tissue specimen collected from a patient with benign prostate hyperplasia undergoing TURP.

A disease condition is primarily marked by a molecular signature which might arise as a result of differential regulation of proteins and/or post-translational modifications of proteins. In this context, mass spectrometry based imaging approach might significantly contribute to understand the disease condition in a better way. Mass of a molecule is its fundamental property. Therefore, mass spectrometry based imaging approach might enable to distinguish between tissues in healthy and disease states based on the differential molecular distribution of ions on the tissue sections. For investigation of marker molecules of various disease conditions, mass spectrometry based proteomics approach is popular among researchers. However, in tissue proteomics, homogenization of tissues results in the loss of spatial heterogeneity of the distribution of molecules across tissue sections. However, mass spectrometry based imaging approach, the two-dimensional heterogeneity of molecular distribution is retained, which results in higher degree of confidence in the investigation of molecular signatures associated with disease states.

In IMS, the molecular images are constructed by using a cartesian coordinate system, for a given mass-to-charge (m/z) signal where the data are represented by an x/y coordinate set and by ‘i’, the intensity of the signal proportional to the abundance of the corresponding molecule. The mass spectrometric molecular images are visualized with different colored codes. A full imaging MS dataset consists of the information in a 2+n dimensional space (x/y/i_n_), where n is the number of m/z signals detected during the course of MS data acquisition. An intensity plot or image is constructed for each m/z value [9].

In the present study, we observed ribonuclease T2, vinculin, isoform 2 of tropomyosin alpha-3 chain and mitochondrial citrate synthase proteins to be expressed across a prostate tissue section with BPH.

## Conclusions

In the present study, MALDI mass spectrometry based tissue imaging platform was used to image the distribution of multiple proteins on human prostate tissues which were diagnosed with cancer. We observed the expression of ribonuclease T2, vinculin, isoform 2 of tropomyosin alpha-3 chain and mitochondrial citrate synthase proteins across the BPH tissue sections.

## Data Availability

Data will be available upon acceptance

## Acknowledgement

We acknowledge all the volunteers for providing tissue samples for the study. We acknowledge Rajiv Gandhi University of Health Sciences, Bangalore, for funding the study and Department of Science and Technology (Nanomission), Govt. of India, for funding mass spectrometry facility at St. John’s Research Institute.

## References

1. Carvalhal, G.F., Smith, D.S., Mager, D.E., Ramos, C., Catalona, W.J.: Digital rectal examination for detecting prostate cancer at prostate specific antigen levels of 4 ng./ml. or less. The Journal of Urology. 161, 835–839 (1999).

2. Mahon, S.M.: Screening for prostate cancer: Informing men about their options. Clinical Journal of Oncology Nursing. 9, 625–627 (2005).

3. Carter, H.B., Hamper, U.M., Sheth, S., Sanders, R.C., Epstein, J.I., Walsh, P.C.: Evaluation of transrectal ultrasound in the early detection of prostate cancer. The Journal of Urology. 142, 1008–1010 (1989).

4. Botchorishvili, G., Matikainen, M.P., Lilja, H.: Early prostate-specific antigen changes and the diagnosis and prognosis of prostate cancer. Current Opinion in Urology. 19, 221–226 (2009).

5. Thompson, I.M., Pauler, D.K., Goodman, P.J., Tangen, C.M., Lucia, M.S., Parnes, H.L., Minasian, L.M., Ford, L.G., Lippman, S.M., Crawford, E.D., Crowley, J.J., Coltman, C.A.: Prevalence of prostate cancer among men with a prostate-specific antigen level ≤4.0 ng per milliliter. New England Journal of Medicine. 350, 2239–2246 (2004).

6. Schwamborn, K.: Imaging mass spectrometry in biomarker discovery and validation. Journal of Proteomics. 75, 4990–4998 (2012).

7. Schwamborn, K., Caprioli, R.M.: Molecular imaging by mass spectrometry-looking beyond classical histology. Nature Reviews Cancer. 10, 639–646 (2010).

8. Chan, J.K.C.: The wonderful colors of the hematoxylin–eosin stain in diagnostic surgical pathology. International Journal of Surgical Pathology. 22, 12–32 (2014).

9. Chaurand, P.: Imaging mass spectrometry of thin tissue sections: A decade of collective efforts. Journal of Proteomics. 75, 4883–4892 (2012).

